# Urinary Creatine Riboside Complements PSA to Improve Disease Detection in the Diagnostic Gray Zone of Prostate Cancer

**DOI:** 10.64898/2026.06.16.26355797

**Authors:** Daxesh P. Patel, Ashlie Santaliz Casiano, Leila Toulabi, Mohammed Khan, Tiffany H. Dorsey, Ewy A. Mathé, Curtis C. Harris, Xin Wei Wang, Stefan Ambs

## Abstract

Circulating prostate-specific antigen (PSA) discriminates poorly in the diagnostic gray zone (3.0–9.99 ng/mL), where ~75% of biopsies yield no clinically significant prostate cancer (PCa). We evaluated whether urinary creatine riboside (CR), a tumor-derived metabolite excreted through the prostatic urethra, complements PSA for gray-zone detection and independently predicts prostate-cancer-specific mortality (PCSM). In the NCI-Maryland PCa Case-Control Study (951 cases, 962 controls; 47.6% African American men; median follow-up 11.5 years), urinary CR was quantified by UPLC-MS/MS. Within the PSA gray zone (*n* = 668), urinary CR was complementary to PSA, with markedly higher single-marker discrimination than PSA (AUC 0.93, 95% CI 0.88–0.98 vs 0.77, 0.66–0.89) and additive when combined (ΔAUC +0.17, *p* < 0.001; 91.4% sensitivity at 80% specificity). After adjustment for 11 clinical and sociodemographic covariates, urinary CR independently predicted PCSM complementary to PSA (Fine–Gray SHR 1.72, 1.35–2.19 for CR; 1.35, 1.08–1.68 for PSA; Harrell’s C 0.85 for CR + PSA vs 0.77 for PSA alone), with strongest signal in African American men (SHR 2.43, 1.57– 3.75 for CR). We conclude that urinary CR is a candidate non-invasive biomarker complementary to PSA — improving gray-zone triage and predicting PCSM; prospective validation in biopsy-referred cohorts is warranted.

## Main text

Each year, approximately one million U.S. men undergo prostate biopsy based on a serum PSA in the 3–10 ng/mL gray zone range, yet only about one in four is diagnosed with clinically significant prostate cancer (PCa) — making improved triage a pressing unmet need. PCa disproportionately affects men of African descent, who experience higher incidence and prostate cancer-specific mortality (PCSM) [1]. Today PCa detection still depends on serum PSA, whose specificity at conventional thresholds is approximately 30%, generating substantial negative-biopsy rates and overdiagnosis [2, 3]. Existing alternatives (4Kscore, Prostate Health Index, PCA3, SelectMDx) reach AUCs of 0.70–0.82 and require venipuncture, post-Digital Rectal Exam (DRE) collection, or prior biopsy [4]. No widely adopted biomarker provides accurate, non-invasive discrimination within the PSA gray zone (3.0–9.99 ng/mL). Our group defined CR as a tumor-derived metabolite of urea-cycle dysregulation [5, 6]. We tested whether urinary CR complements PSA in the gray zone and predicts PCSM.

We analyzed 951 cases and 962 frequency-matched population controls [47.6% African American (AA) men] from the NCI-Maryland PCa Case-Control Study, recruited 2005–2015; survival follow-up extended to December 2022 (median 11.5 years; Table S1) [7, 8]. Voided urine was collected at enrollment; urinary CR was quantified by UPLC-MS/MS and normalized to urinary creatinine [7] (see Supplementary Methods). Diagnostic discrimination used 10-fold cross-validated ROC with paired-bootstrap inference for ΔAUC in the gray zone; 5,000-iteration bootstrap confidence intervals (CIs) were calculated. Mortality was modeled by Cox cause-specific and Fine–Gray subdistribution-hazard regression, fully adjusted for age, BMI, race, treatment (4-category), National Comprehensive Cancer Network (NCCN) PCa risk score, family history, smoking, aspirin, diabetes, education, and individual household income [8]; analytical *n* are in Table S2. Urinary CR and serum PSA were log□-transformed and modeled as continuous exposures.

Full-cohort urinary CR distributions and AUCs (Figure 1A, 1B) reflect by-design case–control separation, not clinical performance. CR and PSA performed similarly. Within the PSA gray zone (*n* = 668), urinary CR was complementary to PSA, with markedly higher single-marker discrimination (AUC 0.93, 95% CI 0.88–0.98 for CR vs 0.77, 0.66–0.89 for PSA; Figure 1C, 1D). At 80% specificity, urinary CR achieved 91.4% sensitivity (Table S3). The combined CR + PSA model achieved AUC 0.94 (0.89–0.99), with ΔAUC vs PSA alone +0.17 (95% CI 0.07– 0.24, *p*<0.001) and Integrated Discrimination Improvement +0.23 (0.16–0.31). Discrimination was strongest in AA men (gray-zone AUC 0.95; Table S4); the case–control distribution at the Youden-optimal threshold of 1.54 µM (Figure 1C) showed near-complete separation.

**Figure 1.**
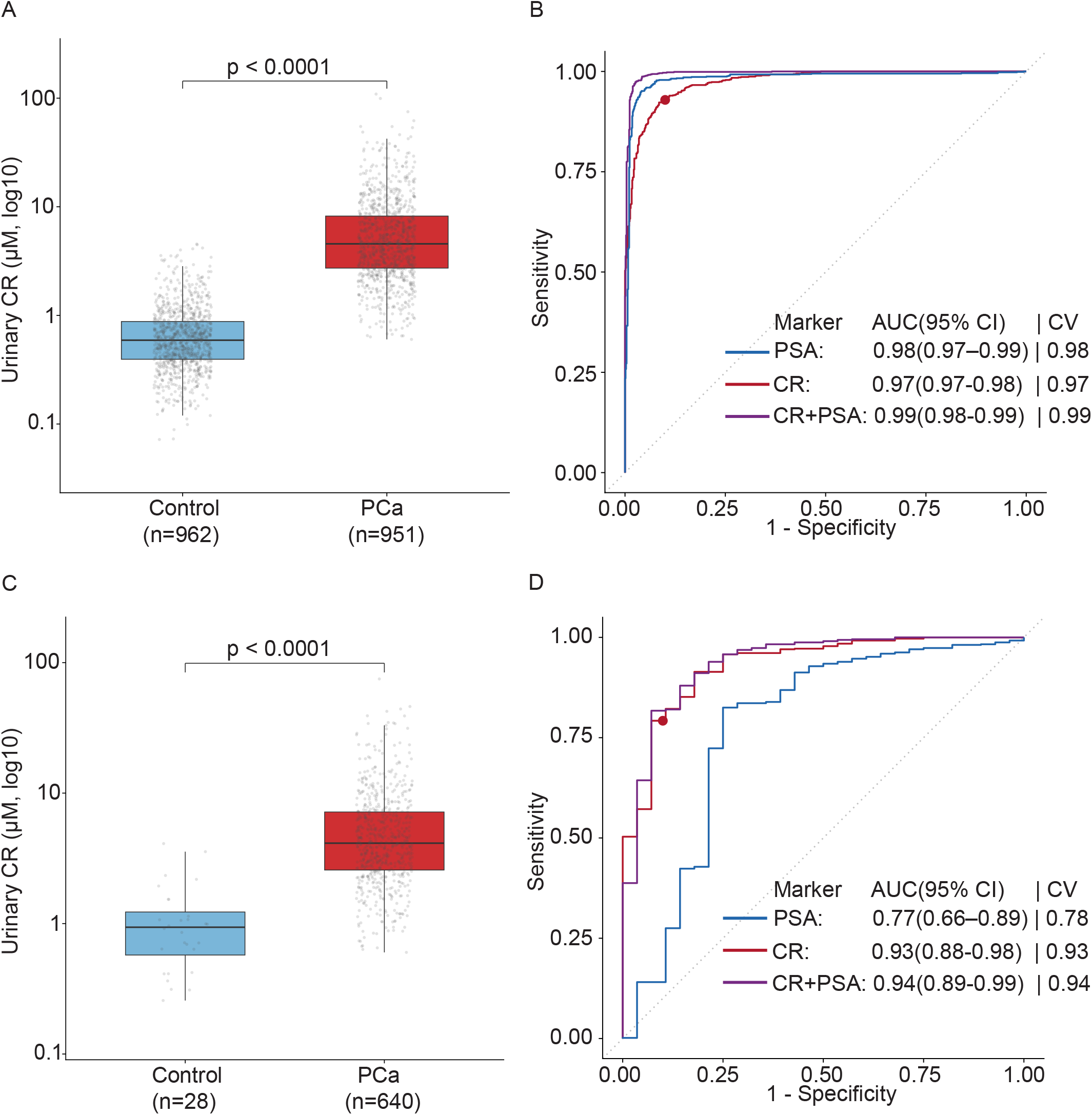
Diagnostic performance of urinary creatine riboside for prostate cancer detection in the full cohort and PSA gray zone. (A) Distribution of urinary CR concentrations (µM, log10) in the full cohort (controls n=962; PCa n=951). (B) ROC curves for PSA, urinary CR, and combined CR+PSA in the full cohort; the red point marks the optimal CR threshold (maximum Youden index). (C) Distribution of urinary CR concentrations in the PSA gray zone subcohort (PSA 3.0–9.99 ng/mL; controls n=28; PCa n=640). (D) ROC curves for the three markers in the PSA gray zone; red point as in B. Distributions compared by Mann–Whitney U test; AUCs estimated by 10-fold cross-validated logistic regression, with ΔAUC compared by paired-bootstrap inference. AUC, area under the curve; CI, confidence interval; CR, creatine riboside; CV, cross-validated; PCa, prostate cancer; PSA, prostate-specific antigen; ROC, receiver operating characteristic.

Kaplan–Meier curves by urinary CR tertile separated for PCSM (Figure 2A; log-rank p<0.0001). Adjusted mortality models used the survival-analysis cohort (n = 854; Supplementary Table 2). In the fully adjusted PCSM model (Figure 2B; Figure S1, Table S5), urinary CR was independently prognostic and complementary to PSA: cause-specific HR 1.71 (1.37–2.15) and Fine–Gray SHR 1.72 (1.35–2.19) for CR, vs 1.36 (1.15–1.62) and 1.35 (1.08–1.68) for PSA. Harrell’s C was 0.85 (0.78–0.90) for CR + PSA vs 0.77 (0.70–0.84) for PSA alone (ΔC +0.07). Effects were largest in AA men (CR SHR 2.43, 1.57–3.75; Figure S2). Estimates were analogous in a sensitivity model excluding socioeconomic covariates (Table S5).

**Figure 2.**
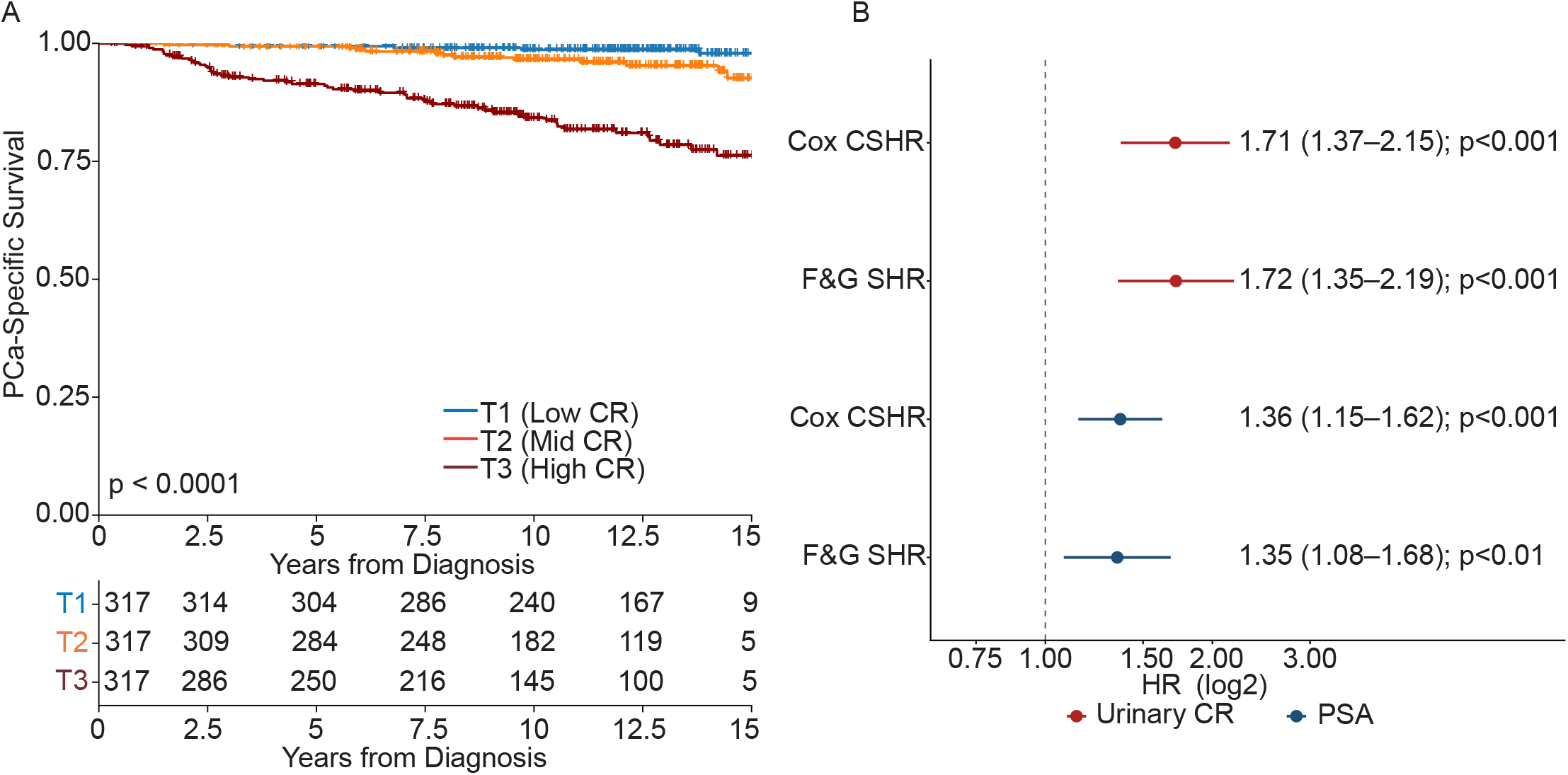
Urinary creatine riboside predicts prostate cancer–specific mortality. (A) Kaplan– Meier estimates of prostate cancer–specific survival stratified by urinary CR tertile (T1, <3.19 μM; T2, 3.19–6.56 μM; T3, >6.56 μM; n=317 per tertile; 72 PCa-specific deaths; median follow-up 11.5 y, IQR 8.1–14.3), compared by log-rank test. (B) Forest plot of adjusted hazard ratios for PCa-specific mortality per 2-fold (log□) increase in urinary CR and PSA (n=854 with complete covariate data), estimated by Cox cause-specific hazard models (CSHR) and Fine–Gray subdistribution hazard models (SHR) accounting for non-cancer death as a competing risk. All multivariable models were adjusted for age, BMI, race, treatment (surgery [ref]/radiotherapy/hormone/multimodal), NCCN risk category (low [ref]/intermediate/high–very high/regional–metastatic), family history of prostate cancer, smoking status, aspirin use, diabetes, education, and individual household income. Abbreviations: BMI, body mass index; CI, confidence interval; CR, creatine riboside; CSHR, cause-specific hazard ratio; F&G, Fine– Gray; HR, hazard ratio; IQR, interquartile range; NCCN, National Comprehensive Cancer Network; PCa, prostate cancer; PSA, prostate-specific antigen; SHR, subdistribution hazard ratio.

The unadjusted urinary CR concentration by AJCC stage (Figure S3) showed stable median CR across stages I–III (3.5–4.7 µM) with a sharp 2.7-fold rise at stage IV (11.7 µM; Kruskal–Wallis *p*<0.0001). For NCCN risk scores, multinomial regression (referent: low NCCN risk) validated this pattern: urinary CR did not differentiate intermediate (RRR 1.01, 0.86–1.19) or high/very-high-risk local disease (RRR 1.10, 0.90–1.34), but was strongly associated with regional/metastatic disease (RRR 1.83, 1.32–2.55; Figure S4 and Table S6). PSA, by contrast, increased monotonically across all NCCN categories. This pattern was confirmed in stage-stratified Cox models (Figure S5). Selective elevation in advanced disease supports urinary CR as a marker of aggressive and metastatic biology rather than tumor burden. As shown in Table S6, PSA showed a stronger monotonic dose-response across NCCN risk categories than urinary CR, indicating PSA is the better readout of NCCN risk.

These findings indicate that PSA tracks tumor burden across the disease spectrum, whereas urinary CR specifically reflects advanced/metastatic biology — most likely arginine auxotrophy and urea-cycle dysregulation in metastatic clones, a metabolic vulnerability being targeted therapeutically with arginine-depleting agents (ADI-PEG20) in clinical trials [5, 6, 9]. This biological complementarity — PSA as a burden marker, urinary CR as an aggressive biology marker — provides the rationale for the additive performance of CR + PSA in the gray zone. Among urinary alternatives in clinical use or under investigation (PCA3, SelectMDx), reported AUCs are 0.70–0.82 [4]; the gray-zone AUC of 0.93 observed here for urinary CR — measured without post-DRE collection — represents a substantial improvement, although this comparison is across studies. Urinary PSA has been proposed as more specific than serum PSA in the gray zone [10] but is not in routine practice; urinary CR offers a different biological signal — tumor metabolism rather than glandular leakage — and is complementary. That urinary CR carries its largest prognostic signal in AA men — the subgroup with the highest PCa mortality — suggests CR captures disease biology that PSA-based stratification underdetects in this population, warranting dedicated validation in AA-enriched cohorts.

Limitations warrant emphasis. The exploratory and retrospective case-control design cannot confirm clinical utility for biopsy decision-making; prospective validation in biopsy-referred populations is required. Intra-prostatic tumor location and volume were unavailable; whether tumor proximity to the urethra modulates urinary CR warrants study, although selective elevation only at stage IV argues against simple volume effects. Mass-spectrometry quantification is not yet point-of-care. Full-cohort AUCs (≥0.97) reflect case–control separation rather than clinical performance; the gray-zone AUC of 0.93 is a clinically realistic estimate. Prospective validation should evaluate urinary CR within MRI-stratified pathways (PI-RADS/mpMRI) to define its incremental value over imaging-based triage. Urinary CR is a non-invasive, dual-utility biomarker that complements serum PSA — improving triage in the diagnostic gray zone and providing independent prognostic stratification for PCSM, with the strongest effect in AA men — and warrants prospective validation.

## Author contributions

Daxesh P. Patel had full access to all the data in the study and takes responsibility for the integrity of the data and the accuracy of the data analysis.

Study concept and design: Patel, Harris, Wang, Ambs. Acquisition of data: Patel, Santaliz Casiano, Toulabi, Dorsey. Analysis and interpretation of data: Patel, Khan.

Drafting of the manuscript: Patel.

Critical revision of the manuscript for important intellectual content: Mathe, Harris, Wang, Ambs.

Statistical analysis: Patel, Khan. Obtaining funding: Harris.

Administrative, technical, or material support: Santaliz Casiano, Toulabi, Dorsey. Supervision: Wang, Ambs.

Other: None.

## Data Availability

All data produced in the present study are available upon reasonable request to the authors. Individual-level data from the NCI-Maryland Prostate Cancer Case-Control Study are not publicly deposited due to participant privacy protections governing this NIH intramural research cohort.

## Use of generative AI and AI-assisted technologies

During the preparation of this manuscript, the authors used Claude Sonnet 4.6 (Anthropic) for grammar and language editing only. After using this tool, the authors reviewed and edited the content as needed and take full responsibility for the content of the publication.

## Acknowledgments

The contributions of the NIH author(s) were made as part of their official duties as NIH federal employees, are in compliance with agency policy requirements, and are considered Works of the United States Government. However, the findings and conclusions presented in this paper are those of the authors and do not necessarily reflect the views of the NIH or the U.S. Department of Health and Human Services.

## References

[1] Rawla P. Epidemiology of Prostate Cancer. World J Oncol. 2019;10:63–89.

[2] Catalona WJ, Partin AW, Sanda MG, Wei JT, Klee GG, Bangma CH, et al. A multicenter study of [-2]pro-prostate specific antigen combined with prostate specific antigen and free prostate specific antigen for prostate cancer detection in the 2.0 to 10.0 ng/ml prostate specific antigen range. J Urol. 2011;185:1650–5.

[3] Pinsky PF, Parnes H. Screening for Prostate Cancer. Reply. N Engl J Med. 2023;389:94.

[4] Kawada T, Shim SR, Quhal F, Rajwa P, Pradere B, Yanagisawa T, et al. Diagnostic Accuracy of Liquid Biomarkers for Clinically Significant Prostate Cancer Detection: A Systematic Review and Diagnostic Meta-analysis of Multiple Thresholds. Eur Urol Oncol. 2024;7:649–62.

[5] Mathe EA, Patterson AD, Haznadar M, Manna SK, Krausz KW, Bowman ED, et al. Noninvasive urinary metabolomic profiling identifies diagnostic and prognostic markers in lung cancer. Cancer Res. 2014;74:3259–70.

[6] Parker AL, Toulabi L, Oike T, Kanke Y, Patel D, Tada T, et al. Creatine riboside is a cancer cell-derived metabolite associated with arginine auxotrophy. J Clin Invest. 2022;132.

[7] Patel DP, Pauly GT, Tada T, Parker AL, Toulabi L, Kanke Y, et al. Improved detection and precise relative quantification of the urinary cancer metabolite biomarkers - Creatine riboside, creatinine riboside, creatine and creatinine by UPLC-ESI-MS/MS: Application to the NCI-Maryland cohort population controls and lung cancer cases. J Pharm Biomed Anal. 2020;191:113596.

[8] Pichardo MS, Minas TZ, Pichardo CM, Bailey-Whyte M, Tang W, Dorsey TH, et al. Association of Neighborhood Deprivation With Prostate Cancer and Immune Markers in African American and European American Men. JAMA Netw Open. 2023;6:e2251745.

[9] Patel R, Ford CA, Rodgers L, Rushworth LK, Fleming J, Mui E, et al. Cyclocreatine Suppresses Creatine Metabolism and Impairs Prostate Cancer Progression. Cancer Res. 2022;82:2565–75.

[10] Bolduc S, Lacombe L, Naud A, Gregoire M, Fradet Y, Tremblay RR. Urinary PSA: a potential useful marker when serum PSA is between 2.5 ng/mL and 10 ng/mL. Can Urol Assoc J. 2007;1:377–81.

