## Supplement_Material for "Urinary Creatine Riboside Complements PSA to Improve Disease Detection in the Diagnostic Gray Zone of Prostate Cancer"

**Contents**

**Supplementary Methods**

1. Study cohort, recruitment, and eligibility criteria

2. Urinary CR quantification by LC-MS/MS

3. Treatment details

4. Statistical methods

**Supplementary Tables**

Supplementary Table 1. Baseline characteristics of the NCI-Maryland Prostate Cancer Case-Control Study

Supplementary Table 2. Analytical cohort characteristics for survival analyses

Supplementary Table 3. Sensitivity of urinary CR at fixed specificity thresholds

Supplementary Table 4. Diagnostic performance of urinary CR and PSA in the entire NCI-MD cohort and among men in the PSA gray zone

Supplementary Table 5. Association of urinary CR and PSA with prostate-cancer-specific mortality (Cox cause-specific and Fine–Gray subdistribution-hazard regression)

Supplementary Table 6. Association of urinary CR and PSA with NCCN risk category at diagnosis (multinomial logistic regression)

**Supplementary Figures**

Supplementary Figure 1. Full multivariable forest plots for prostate cancer–specific mortality

Supplementary Figure 2. Race-stratified multivariable hazard ratios for urinary CR and PSA with prostate cancer–specific mortality

Supplementary Figure 3. Urinary CR concentration by clinical risk stratification at diagnosis (NCCN risk category and AJCC clinical stage)

Supplementary Figure 4. Multinomial relative risk ratios (RRRs) for urinary CR across NCCN risk categories, overall and race-stratified

Supplementary Figure 5. Stage-stratified hazard ratios for urinary CR on prostate cancer–specific mortality

**References**

### Supplementary Methods

### 1. Study cohort, recruitment, and eligibility criteria

### *NCI-Maryland Prostate Cancer Case-Control Study*

### The NCI-Maryland (NCI-MD) Prostate Case-Control Cancer (PCa) Study is a population-based study of PCa among self-identified African American (AA; Black) and European American (EA; White) men, conducted between January 1, 2005 and December 31, 2015 [1, 2]. Eligible cases were men aged 40–90 years with histologically confirmed incident PCa diagnosed within 12 months of enrollment, recruited from the Baltimore Veterans Affairs Medical Center and the University of Maryland Medical Center. Population-based controls were identified from the Maryland Department of Motor Vehicle Administration database and frequency-matched to cases by 5-year age group and self-reported race; controls had no prior history of PCa. Men with a prior history of cancer (other than non-melanoma skin cancer) were excluded.

### At enrollment, trained interviewers administered standardized questionnaires capturing demographics, medical history, medication use (including aspirin), family history of prostate cancer, smoking status, and anthropometric measurements. Voided urine specimens were collected at enrollment and stored at −80 °C until analysis. Clinical data, including TNM stage, Final Gleason score, and serum prostate-specific antigen (PSA) at diagnosis, were extracted from medical records. National Comprehensive Cancer Network (NCCN) risk categories [3] were assigned and collapsed into four groups: low; intermediate (favorable + unfavorable); high or very high; and regional or metastatic, as previously described [1, 2]. National Death Index linkage was performed for vital-status follow-up through December 31, 2022.

### After exclusions for missing baseline address (n = 86), prevalent (non-incident) cases (n = 131), and missing covariates (n = 1), the full cohort comprised 951 cases and 962 controls (overall 47.6% AA). Serum PSA values were available for 942 cases and 779 controls.

### *Cohort characteristics*

### Median follow-up among cases was 11.5 years (survival-analysis cohort). Within the full case cohort (n = 951; Supplementary Table 1), 72 prostate-cancer-specific deaths (PCSM) and 268 competing-cause deaths were recorded; in the complete-case Fine–Gray PCSM model (n = 854; Supplementary Table 2) there were 63 PCa-specific deaths and 238 competing-cause deaths. Compared with EA men, AA men were younger (mean age 62.2 vs 64.3 years), more likely to report household income <$30,000 (56.6% vs 24.7%) and current smoking (35.4% vs 16.3%) and had a higher prevalence of diabetes (31.1% vs 18.9%; Supplementary Table 1). NCCN risk distribution was comparable between groups; regional/metastatic disease was present at diagnosis in 6.3% of AA and 4.5% of EA men. Median urinary creatine riboside (CR) was higher in AA men (5.39 µM, IQR 3.19–9.62) than in EA men (3.75 µM, IQR 2.34–7.00), whereas median serum PSA was comparable between groups (6.98 vs 5.99 ng/mL).

### *Ethics approval*

### The study was approved by the Institutional Review Boards of the National Cancer Institute (protocol #05-C-N021) and the University of Maryland (protocol #0298229). All participants provided written informed consent.

### 2. Urinary CR quantification by LC-MS/MS

### Urinary CR and creatinine were simultaneously quantified by ultra-performance liquid chromatography coupled to electrospray-ionization tandem mass spectrometry (UPLC-ESI-MS/MS), as previously described [4]. Analysis was performed on a Waters Acquity UPLC system coupled to a Waters Xevo TQ-S micro triple-quadrupole mass spectrometer operating in positive ESI mode. Chromatographic separation was achieved on a HILIC Acquity UPLC BEH Amide column (1.7 µm, 2.1 × 50 mm) using a gradient mobile phase of 10 mM ammonium acetate in 90% acetonitrile (A; pH 9.0) and 10 mM ammonium acetate in 10% acetonitrile (B; pH 9.0). Detection used multiple reaction monitoring (MRM). Transitions monitored were CR (m/z 264.1 → 132.1), creatinine (m/z 114.0 → 85.8), and the stable-isotope-labelled internal standard creatine riboside-¹³C,¹⁵N₂ (m/z 267.1 → 134.9). Total analytical run time was 11.0 min. Full chromatographic gradient and mass-spectrometric parameters are detailed in Patel et al. [4].

### *Urine sample preparation*

### Urine (10 µL) was diluted with acetonitrile/water/methanol (65:30:5, v/v/v) and vortexed; 80 µL was transferred and combined with 80 µL of 75% acetonitrile containing 5 µM creatine riboside-¹³C,¹⁵N₂ internal standard. Samples were vortexed, centrifuged at 20,000 × g for 10 min at 4 °C, and the supernatant was transferred to LC-MS/MS vials for injection (5 µL). Urinary CR concentrations were normalized to creatinine measured in the same analytical run to account for variation in urine dilution.

### *Method validation*

### The method was validated in accordance with US FDA bioanalytical guidelines [5]: linear range 4.50–10,000 nM (R² ≥ 0.9936); intra- and inter-day precision (%CV) 3.1–9.5%; accuracy 96–105%; lower limit of quantification 1.5 nM.

### 3. Treatment details

### Primary definitive treatment after diagnosis was ascertained from study questionnaires and medical records and grouped into four categories for adjusted survival analyses: radical prostatectomy alone (reference); radiation therapy alone (external-beam radiotherapy, brachytherapy, or both); hormone or androgen-deprivation therapy alone; and multimodal therapy (any combination of surgery, radiation, hormonal therapy, and/or chemotherapy). Cases without a recorded definitive treatment (n = 119) were retained within the multimodal group as an “untreated” stratum for inclusive sensitivity analyses; estimates were materially unchanged when these cases were excluded.

### 4. Statistical methods

### *Marker modelling and diagnostic accuracy*

### Urinary CR and serum PSA were log₂-transformed and modelled as continuous exposures; hazard and risk ratios correspond to a 2-fold (i.e., doubling) increase in marker concentration unless otherwise stated. Diagnostic accuracy was evaluated using receiver-operating-characteristic (ROC) curve analysis. Areas under the curve (AUCs) and 95% confidence intervals (CIs) were estimated by 10-fold stratified cross-validation, with logistic-regression models trained on 90% of the data and evaluated on the held-out 10% within each fold; CIs were derived by 5,000-iteration bootstrap resampling. Within the PSA gray zone (3.0–9.99 ng/mL; n = 668), urinary CR alone, PSA alone, and the combined CR + PSA logistic-regression model were compared using paired-bootstrap inference for ΔAUC. Sensitivity at fixed specificity thresholds was computed at 80%, 85%, 90%, and 95%, and at the Youden-optimal cut-point.

### *Risk and survival models*

### Multinomial logistic regression was used to estimate relative risk ratios (RRRs) for the association of urinary CR and PSA with NCCN risk category at diagnosis (referent, low). For prostate-cancer-specific mortality, Cox cause-specific models (censoring competing-cause deaths) and Fine–Gray subdistribution-hazard models (treating deaths from non-prostate-cancer causes as competing events) were both fit. The proportional-hazards assumption was verified via scaled Schoenfeld residuals. Median follow-up was estimated by reverse Kaplan–Meier (full cohort, 11.5 yr; PCa-specific mortality, 11.8 yr).

### *Covariate adjustment*

***Primary model.*** The primary fully adjusted model, used for all main-text estimates and following the framework of Pichardo et al. [1], included age at study entry (continuous), body mass index (kg/m², continuous), self-reported race (omitted in race-stratified analyses), treatment (4-category, as defined in Supplementary Methods 3), family history of prostate cancer (yes/no), smoking history (never/former/current), aspirin use (yes/no), diabetes (yes/no), education (≤ high school, Some college, college graduate, postgraduate / professional, technical school), and individual household income (<$30,000, $30,000–$60,000, $60,000–$90,000, >$90,000). For mortality models, NCCN risk category was additionally included.

***Sensitivity model.*** As a sensitivity analysis, a reduced model excluding education and individual household income but otherwise identical to the primary model was fit (the primary fully adjusted model is referred to as Model 1 and the reduced sensitivity model as Model 2 in the supplementary tables). This sensitivity model evaluates whether the urinary CR signal is stable to the inclusion or exclusion of socioeconomic covariates and aids interpretation against the broader literature, in which some studies do not include detailed socioeconomic adjustment. Estimates from the primary and sensitivity models were materially indistinguishable across all analyses.

***Analytical sample size.*** Because all models use complete-case analysis, the analytical sample is larger for the sensitivity model than for the primary model. Education and individual household income carry the most item missingness among the covariates, so cases missing one of these socioeconomic variables but complete on all others are excluded from the primary model yet retained in the sensitivity model. Dropping these two covariates therefore adds those cases back, increasing the analytical n rather than reducing it. For prostate-cancer-specific mortality, the primary model included n = 854 cases (AA n = 420; EA n = 434) and the corresponding sensitivity model n = 930 cases (AA n = 466; EA n = 464), an increase of 76 cases (Supplementary Table 2). Analytical n also differs modestly across endpoints because the Fine–Gray PCSM model and the multinomial NCCN model require slightly different sets of non-missing covariates.

### Stratification and software

### Analyses were stratified by self-reported race (AA, EA) in addition to the unstratified analysis. Two-sided *p* < 0.05 was considered statistically significant. All analyses were performed in R 4.3 using the survival, cmprsk, nnet, and pROC packages.

### Supplementary Tables

### Supplementary Table 1. Baseline characteristics of the NCI-Maryland Prostate Cancer Case-Control Study

| **Characteristic** | **Cases (n = 951)** | **Controls (n = 962)** | ***p* value** |
| --- | --- | --- | --- |
| ***Demographic characteristics*** |  |  |  |
| Age at enrollment, years, median (IQR) | 63 (58–68) | 65 (60–72) | <0.001 |
| **Age category, n (%)** |  |  | <0.001 |
| 40–54 | 123 (12.9) | 57 (5.9) |  |
| 55–64 | 437 (46.0) | 395 (41.1) |  |
| 65–74 | 313 (32.9) | 347 (36.1) |  |
| ≥75 | 78 (8.2) | 163 (16.9) |  |
| **Self-reported race, n (%)** |  |  | 0.013 |
| AA | 480 (50.5) | 430 (44.7) |  |
| EA | 471 (49.5) | 532 (55.3) |  |
| ***Anthropometric*** |  |  |  |
| Body mass index, kg/m², median (IQR) | 27.61 (25.04–30.68) | 27.76 (24.96–31.19) | 0.508 |
| **BMI category, n (%)** |  |  | 0.546 |
| Normal (<25) | 235 (24.9) | 245 (25.5) |  |
| Overweight (25–29.9) | 424 (45.0) | 410 (42.6) |  |
| Obese (≥30) | 283 (30.0) | 307 (31.9) |  |
| ***Behavioral*** |  |  |  |
| **Smoking status, n (%)** |  |  | <0.001 |
| Never | 316 (33.6) | 392 (40.7) |  |
| Former | 381 (40.5) | 438 (45.5) |  |
| Current | 244 (25.9) | 132 (13.7) |  |
| ***Clinical history*** |  |  |  |
| Family history of PCa, n (%) | 101 (10.7) | 55 (5.7) | <0.001 |
| Diabetes mellitus, n (%) | 237 (25.0) | 214 (22.2) | 0.169 |
| Aspirin use, n (%) | 463 (49.5) | 454 (57.0) | 0.002 |
| ***Socioeconomics*** |  |  |  |
| **Education, n (%)** |  |  | <0.001 |
| ≤ High school | 348 (37.1) | 246 (26.4) |  |
| Some college | 255 (27.2) | 211 (22.7) |  |
| College graduate | 165 (17.6) | 242 (26.0) |  |
| Postgraduate / professional | 135 (14.4) | 211 (22.7) |  |
| Technical school | 35 (3.7) | 21 (2.3) |  |
| **Individual household income, n (%)** |  |  | <0.001 |
| <$30,000 | 351 (40.5) | 145 (16.9) |  |
| $30,000–$60,000 | 202 (23.3) | 228 (26.6) |  |
| $60,000–$90,000 | 137 (15.8) | 211 (24.6) |  |
| >$90,000 | 176 (20.3) | 272 (31.8) |  |
| ***Biomarkers at enrollment*** |  |  |  |
| Serum PSA, ng/mL, median (IQR) | 6.30 (4.80–10.67) | 0.43 (0.23–0.84) | <0.001 |
| **PSA category, ng/mL, n (%)** |  |  | <0.001 |
| <3.0 | 46 (4.8) | 749 (77.9) |  |
| 3.0–9.99 | 640 (67.3) | 28 (2.9) |  |
| ≥10 | 256 (26.9) | 2 (0.2) |  |
| Urinary CR, µM, median (IQR) | 4.56 (2.73–8.21) | 0.59 (0.39–0.88) | <0.001 |
| PSA in gray zone (3.0–9.99 ng/mL), n (%) | 640 (67.3) | 28 (2.9) | <0.001 |
| PSA, ng/mL, median (IQR) | 5.53 (4.60–6.99) | 3.87 (3.38–4.45) | <0.001 |
| Urinary CR, µM, median (IQR) | 4.13 (2.57–7.17) | 0.94 (0.58–1.24) | <0.001 |
| ***Patient characteristics*** |  |  |  |
| **NCCN risk score category, n (%)** |  | — |  |
| Low | 182 (19.2) |  |  |
| Intermediate | 493 (51.9) |  |  |
| High or very high | 223 (23.5) |  |  |
| Regional or metastatic | 51 (5.4) |  |  |
| **Final Gleason score, n (%)** |  | — |  |
| ≤6 | 393 (41.3) |  |  |
| 7 | 392 (41.2) |  |  |
| ≥8 | 166 (17.5) |  |  |
| **Clinical stage, n (%)** |  | — |  |
| I | 182 (19.1) |  |  |
| II (IIA + IIB) | 636 (66.9) |  |  |
| III | 76 (8.0) |  |  |
| IV | 57 (6.0) |  |  |
| ***Follow-up*** |  |  |  |
| Median follow-up, years (IQR) | 11.5 (8.1–14.3) | — |  |
| PCa-specific deaths, n (%) | 72 (7.6) | — |  |

**Abbreviations.** AA, African American; BMI, body-mass index; CR, creatine riboside; EA, European American; IQR, interquartile range; NCCN, National Comprehensive Cancer Network; PCa, prostate cancer; PCSM, prostate-cancer-specific mortality; PSA, prostate-specific antigen.

**Notes.** Continuous variables compared by Mann–Whitney U test; categorical variables by Pearson χ² test. Tumour-characteristic block reports cases only; em-dashes (—) denote not applicable. Denominators for variables with item missingness exclude missing values; percentages are within-column. The PSA-category sub-block uses the full case (n = 951) and control (n = 962) groups as denominators (missing PSA: 9 cases and 183 controls). Follow-up statistics refer to the full case and control cohort; the **survival-analysis cohort** used in adjusted models comprises 854 cases for PCa-specific mortality (63 PCa-specific deaths; Supplementary Table 2).All values were computed from the source dataset.

**Supplementary Table 2. Survival-analysis cohort characteristics**

|  | **All men** | **AA men** | **EA men** |
| --- | --- | --- | --- |
| **Primary-model analytical n** | 854 | 420 | 434 |
| **Sensitivity-model analytical n** | 930 | 466 | 464 |
| **Median follow-up, years (IQR)** | 11.5 (8.1–14.3) | 10.4 (7.5–13.7) | 12.1 (8.8–14.8) |
| **PCa-specific deaths (primary)** | 63 | 38 | 25 |
| **Competing-cause deaths (primary)** | 238 | 117 | 121 |

**Abbreviations.** AA, African American; EA, European American; NCCN, National Comprehensive Cancer Network; PCa, prostate cancer; PCSM, prostate-cancer-specific mortality.

**Notes.** Differences in analytical n between the sensitivity and primary models reflect item missingness in the education and individual household income variables, which are included in the primary fully adjusted model but excluded from the sensitivity model; because complete-case analysis is used, dropping these two socioeconomic covariates retains cases that are otherwise complete, so the sensitivity-model cohort is larger than the primary-model cohort. The primary-model cohort (N = 854; AA n = 420; EA n = 434) applies to the PCSM Cox cause-specific and Fine–Gray subdistribution-hazard models (Supplementary Table 5) and to the multinomial NCCN model (Supplementary Table 6); minor differences in n across endpoints reflect complete-case requirements that differ between models, because the Fine–Gray PCSM model and the multinomial NCCN model require slightly different sets of non-missing covariates.

**Supplementary Table 3. Sensitivity of urinary CR at fixed specificity thresholds**

| **Cohort** | **Specificity target** | **CR sensitivity** | **CR threshold (µM)** |
| --- | --- | --- | --- |
| **Full NCI-MD (n = 1,913)** | 80% | 97.3% | 1.00 |
|  | 85% | 96.2% | 1.15 |
|  | 90% | 93.7% | 1.40 |
|  | 95% | 87.2% | 1.92 |
|  | Youden optimal | 92.3% (at 92.3% specificity) | 1.54 |
| **Gray zone (n = 668)** | 80% | 91.4% | 1.53 |
|  | 85% | 85.2% | 1.96 |
|  | 90% | 82.2% | 2.18 |
|  | 95% | 57.2% | 3.56 |
|  | Youden optimal | 91.4% (at 82.1% specificity) | 1.54 |

**Abbreviations.** CR, creatine riboside; NCI-MD, National Cancer Institute–Maryland; PSA, prostate-specific antigen.

**Notes.** Sensitivity at clinically interpretable specificity thresholds. The Youden-optimal threshold maximizes sensitivity + specificity − 1.

**Supplementary Table 4. Diagnostic performance of urinary CR and PSA in the full NCI-MD cohort and among men in the PSA gray zone**

***Panel A.*** *Areas under the ROC curve, by cohort and PSA gray-zone subgroup.*

| **Cohort / subgroup** | **n** | **PSA alone AUC (95% CI)** | **Urinary CR alone AUC (95% CI)** | **CR + PSA combined AUC (95% CI)** |
| --- | --- | --- | --- | --- |
| **Full cohort** | 1,721 | 0.98 (0.97–0.99) | 0.97 (0.97–0.98) | 0.99 (0.98–0.99) |
| **AA** | 825 | 0.98 (0.97–0.99) | 0.98 (0.97–0.98) | 0.99 (0.99–1.00) |
| **EA** | 896 | 0.98 (0.97–0.99) | 0.97 (0.96–0.98) | 0.99 (0.99–1.00) |
| **PSA gray zone in full cohort (3.0–9.99 ng/mL)** | **668** | **0.77 (0.66–0.89)** | **0.93 (0.88–0.98)** | **0.94 (0.89–0.99)** |
| **Gray zone — AA** | 321 | 0.85 (0.73–0.95) | 0.95 (0.90–0.99) | 0.96 (0.90–0.99) |
| **Gray zone — EA** | 347 | 0.69 (0.48–0.88) | 0.92 (0.81–0.98) | 0.89 (0.75–0.99) |

***Panel B.*** *Pairwise AUC comparisons within the PSA gray zone (paired bootstrap, n = 668).*

| **Comparison** | **ΔAUC** | **95% CI** | ***p* value** |
| --- | --- | --- | --- |
| CR alone vs PSA alone | +0.16 | +0.06 to +0.26 | 0.001 |
| **CR + PSA vs PSA alone** | **+0.17** | **+0.07 to +0.24** | **<0.001** |
| CR + PSA vs CR alone | −0.01 | −0.04 to +0.02 | 0.78 |

**Abbreviations.** AA, African American; AUC, area under the curve; CI, confidence interval; CR, creatine riboside; EA, European American; NCI-MD, National Cancer Institute–Maryland; PSA, prostate-specific antigen; ROC, receiver operating characteristic.

**Notes.** AUCs estimated by 10-fold stratified cross-validated logistic regression; 95% CIs from 5,000-iteration bootstrap resampling. PSA and urinary CR were log₁₀-transformed for ROC analysis. The Full NCI-MD analytical n = 1,721 reflects exclusion of 192 participants with missing serum PSA at enrollment (9 cases and 183 controls; see Supplementary Table 1). The high full-cohort AUCs (≥0.97) reflect near-complete case–control separation expected in a population-based design in which most controls have low PSA; the gray-zone AUC of 0.93 represents the clinically relevant scenario. Pairwise AUC differences (Panel B) were tested by paired-bootstrap inference for ΔAUC on the same gray-zone participants. Subgroup AUCs are descriptive; the study was not powered for between-subgroup comparison.

**Supplementary Table 5. Association of urinary CR and PSA with prostate-cancer-specific mortality (Cox cause-specific and Fine–Gray subdistribution-hazard regression)**

| **Model / Marker** | **All men (N=854) Primary** | **All men (N=930) Sensitivity** | **AA men (n=420) Primary** | **AA men (n=466) Sensitivity** | **EA men (n=434) Primary** | **EA men (n=464) Sensitivity** |
| --- | --- | --- | --- | --- | --- | --- |
| **Urinary CR — Cox CSHR (95% CI)** | **1.71**  **(1.37–2.15)** | 1.61  (1.33–1.95) | **2.50**  **(1.87–3.35)** | 2.04  (1.55–2.69) | 1.20  (0.85–1.69) | 1.13  (0.83–1.55) |
| **Urinary CR — Fine & Gray SHR (95% CI)** | **1.72**  **(1.35–2.19)** | 1.65  (1.35–2.01) | **2.43**  **(1.57–3.75)** | 2.05  (1.49–2.82) | 1.22  (0.92–1.63) | 1.18  (0.90–1.55) |
| **Serum PSA — Cox CSHR (95% CI)** | 1.36  (1.15–1.62) | 1.32  (1.15–1.53) | 1.56  (1.38–1.77) | 1.52  (1.24–1.86) | 0.77  (0.54–1.10) | 0.85  (0.61–1.18) |
| **Serum PSA — Fine & Gray SHR (95% CI)** | 1.35  (1.08–1.68) | 1.31  (1.10–1.57) | 1.53  (1.20–1.95) | 1.50  (1.24–1.82) | 0.79  (0.39–1.63) | 0.89  (0.47–1.70) |

**Abbreviations.** AA, African American; CI, confidence interval; CR, creatine riboside; CSHR, cause-specific hazard ratio (Cox); EA, European American; NCCN, National Comprehensive Cancer Network; PCa, prostate cancer; PCSM, prostate-cancer-specific mortality; PSA, prostate-specific antigen; SHR, subdistribution hazard ratio (Fine–Gray).

**Notes.** Urinary CR and PSA were log₂-transformed and modelled as continuous exposures; hazard ratios correspond to a 2-fold increase in marker concentration. Cox cause-specific models censor competing-cause deaths; Fine–Gray subdistribution-hazard models treat deaths from non-prostate-cancer causes as competing events. Both models include NCCN risk category, treatment (4 categories), and the remaining covariates listed in Supplementary Methods 4. PCa-specific deaths: 63 among cases with complete covariate data (38 in AA men, 25 in EA men); 238 competing-cause deaths. Bold primary-model estimates for all men correspond to those reported in the abstract and Figure 2B of the main text. Harrell's C-index for discrimination of PCa-specific mortality: PSA alone, 0.77 (95% CI 0.70–0.84); CR alone, 0.83 (0.77–0.89); CR + PSA, 0.85 (0.78–0.90); ΔC (CR + PSA vs PSA alone) +0.07.

**Supplementary Table 6. Association of urinary CR and PSA with NCCN risk category at diagnosis (multinomial logistic regression)**

| **Stratum and risk category (referent: Low)** | **Urinary CR — RRR (95% CI)** | **PSA — RRR (95% CI)** |
| --- | --- | --- |
| ***All men, primary model (n = 854)*** |  |  |
| Intermediate | 1.01 (0.86–1.19) | 2.12 (1.62–2.78) |
| High / Very High | 1.10 (0.90–1.34) | 5.62 (4.02–7.86) |
| Regional / Metastatic | 1.83 (1.32–2.55) | 14.44 (9.04–23.04) |
| ***All men, sensitivity model (n = 930)*** |  |  |
| Intermediate | 1.01 (0.86–1.18) | 2.07 (1.61–2.67) |
| High / Very High | 1.15 (0.96–1.39) | 5.32 (3.89–7.27) |
| Regional / Metastatic | 2.12 (1.55–2.90) | 12.04 (8.03–18.06) |
| ***AA men, primary model (n = 420)*** |  |  |
| Intermediate | 1.10 (0.85–1.42) | 3.51 (1.93–6.39) |
| High / Very High | 1.35 (0.98–1.87) | 13.76 (6.76–27.98) |
| Regional / Metastatic | 2.31 (1.36–3.95) | 43.13 (17.92–103.79) |
| ***AA men, sensitivity model (n = 466)*** |  |  |
| Intermediate | 1.09 (0.85–1.39) | 3.26 (1.92–5.53) |
| High / Very High | 1.40 (1.04–1.89) | 10.14 (5.52–18.63) |
| Regional / Metastatic | 2.42 (1.53–3.85) | 26.86 (12.97–55.65) |
| ***EA men, primary model (n = 434)*** |  |  |
| Intermediate | 0.95 (0.75–1.19) | 1.90 (1.40–2.58) |
| High / Very High | 1.04 (0.80–1.35) | 4.32 (2.88–6.48) |
| Regional / Metastatic | 1.92 (1.13–3.25) | 8.42 (4.40–16.11) |
| ***EA men, sensitivity model (n = 464)*** |  |  |
| Intermediate | 0.97 (0.79–1.20) | 1.81 (1.36–2.41) |
| High / Very High | 1.07 (0.84–1.36) | 4.09 (2.80–5.98) |
| Regional / Metastatic | 2.04 (1.27–3.26) | 8.17 (4.69–14.25) |

**Abbreviations.** AA, African American; CI, confidence interval; CR, creatine riboside; EA, European American; NCCN, National Comprehensive Cancer Network; PCa, prostate cancer; PSA, prostate-specific antigen; RRR, relative risk ratio.

**Notes. Primary model:** fully adjusted for age, body-mass index, self-reported race (unstratified analyses only), treatment, family history of PCa, smoking status, aspirin use, diabetes, education, and individual household income. **Sensitivity model:** reduced model excluding education and individual household income but otherwise identical to the primary model. Urinary CR and PSA were log₂-transformed and modelled as continuous exposures; RRRs correspond to a 2-fold increase in marker concentration. Estimates from the primary and sensitivity models were materially indistinguishable, supporting robustness of the urinary CR signal to socioeconomic adjustment. Bold values indicate the regional/metastatic RRR for urinary CR within each stratum.

### Supplementary Figures

**Supplementary Figure 1. Full multivariable forest plots for prostate cancer–specific mortality (n=854).**
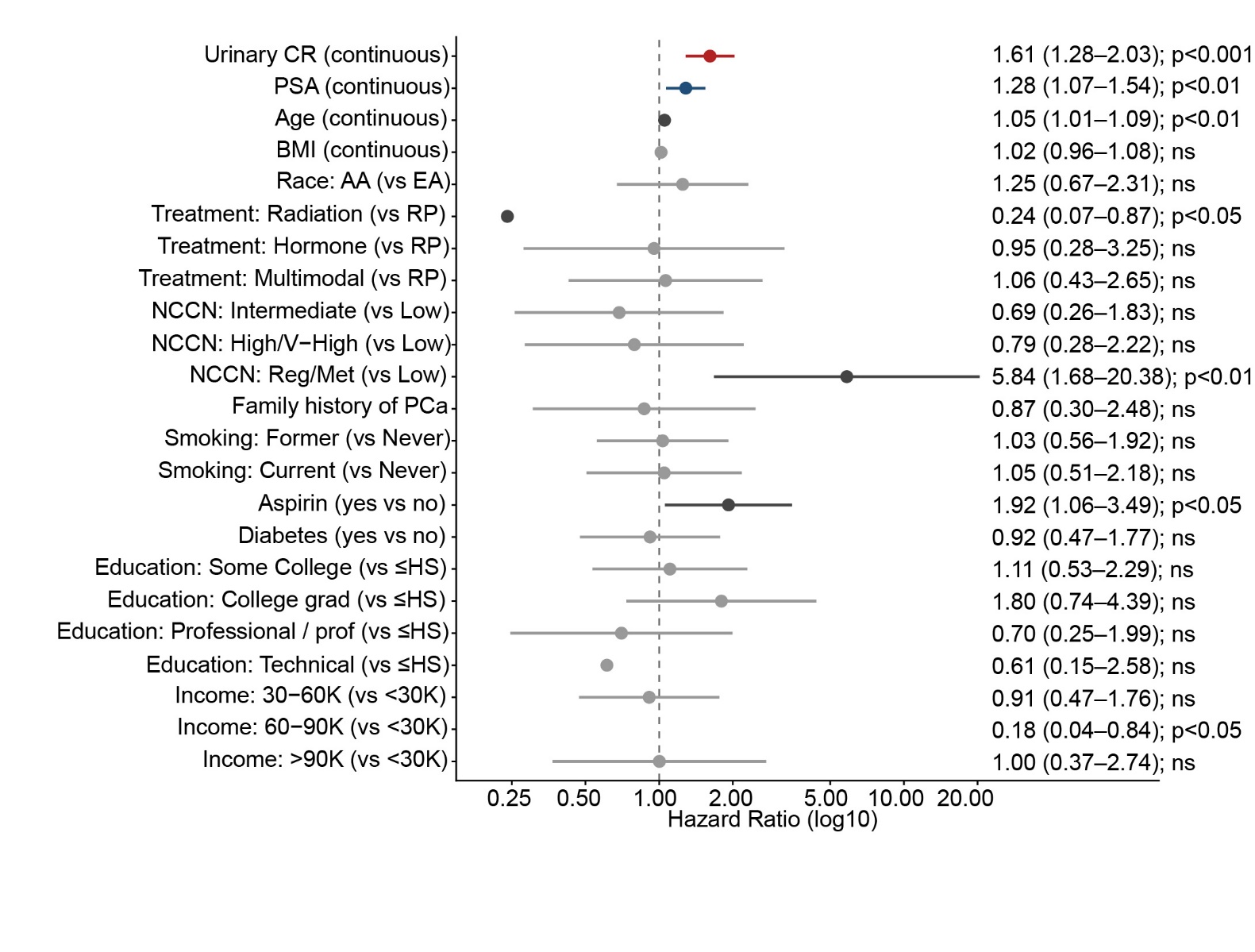

Cox cause-specific hazard model for prostate cancer–specific mortality, accounting for non–prostate cancer death as a competing risk. Adjusted hazard ratios are shown for urinary CR, PSA, and all clinical and sociodemographic covariates. Hazard ratios are plotted on a log₁₀ scale per 2-fold increase in continuous markers (urinary CR and PSA). Points indicate adjusted hazard ratios and horizontal bars represent 95% confidence intervals; the dashed vertical line denotes the null (HR = 1.0). Red = urinary CR; blue = PSA; gray = clinical and sociodemographic covariates. The model was adjusted for age, BMI, race, treatment (radical prostatectomy [reference]/radiotherapy/hormone/multimodal), NCCN risk category, family history of prostate cancer, smoking status, aspirin use, diabetes, education, and individual household income. Abbreviations: AA, African American; BMI, body mass index; CI, confidence interval; CR, creatine riboside; EA, European American; HR, hazard ratio; NCCN, National Comprehensive Cancer Network; PCa, prostate cancer; PSA, prostate-specific antigen; RP, radical prostatectomy.

**Supplementary Figure 2. Race-stratified multivariable hazard ratios for the relationship of urinary CR and PSA with prostate cancer–specific mortality.**

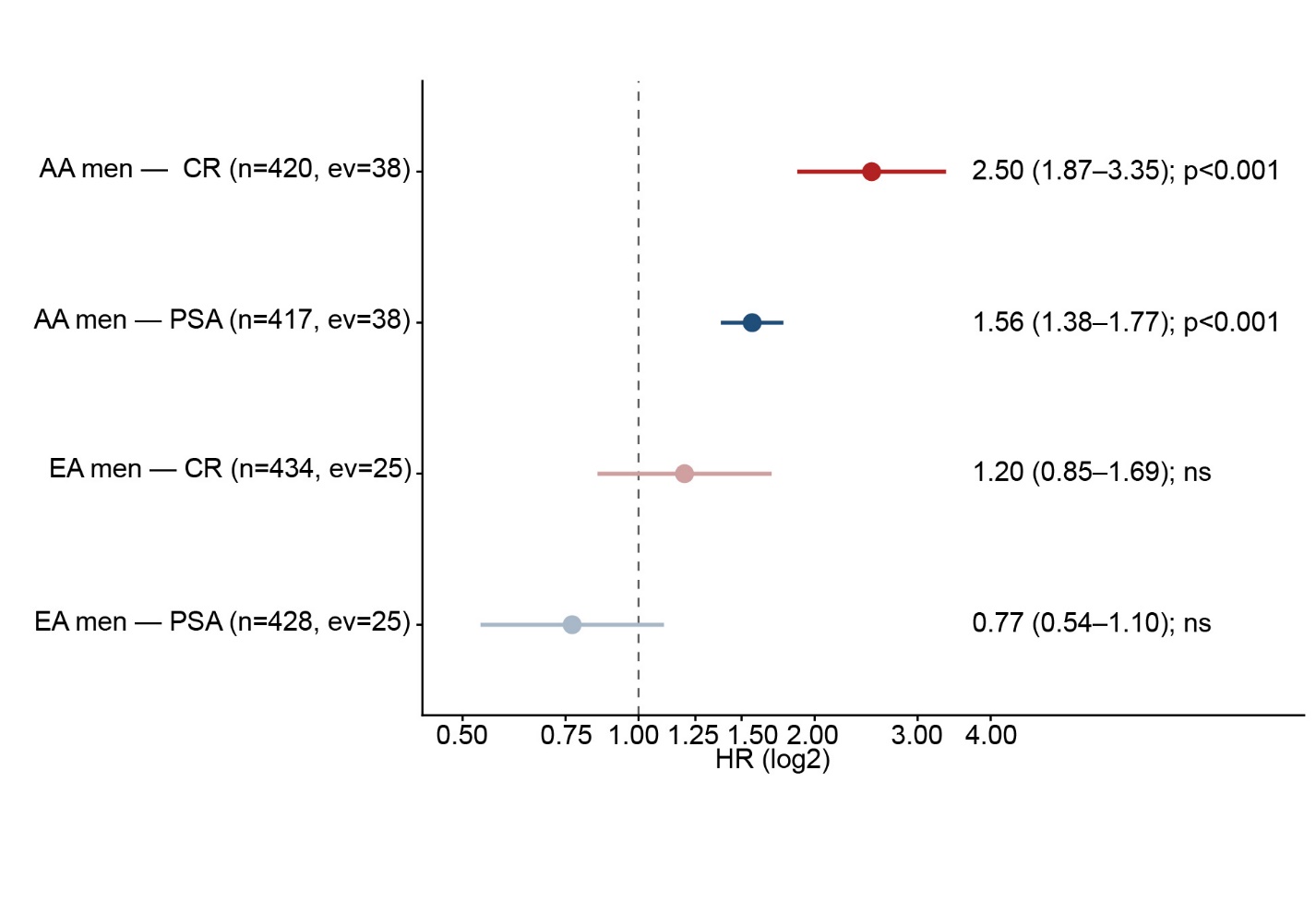

Cox proportional hazards models stratified by self-reported race in AA and EA men, comparing urinary CR and serum PSA as continuous predictors of prostate cancer–specific mortality per 2-fold (log₂) increase in marker concentration. AA men: n=420 for CR and n=417 for PSA (38 PCa-specific deaths). EA men: n=434 for CR and n=428 for PSA (25 PCa-specific deaths). All models were adjusted for age, BMI, treatment modality, NCCN risk category, family history of prostate cancer, smoking status, aspirin use, diabetes, education, and individual household income (race dropped within stratum). Red = urinary CR; blue = PSA. Abbreviations: AA, African American; BMI, body mass index; CI, confidence interval; CR, creatine riboside; EA, European American; HR, hazard ratio; NCCN, National Comprehensive Cancer Network; ns, not significant; PCa, prostate cancer; PSA, prostate-specific antigen.

**Supplementary Figure 3. Urinary CR concentration by clinical risk stratification at diagnosis.**

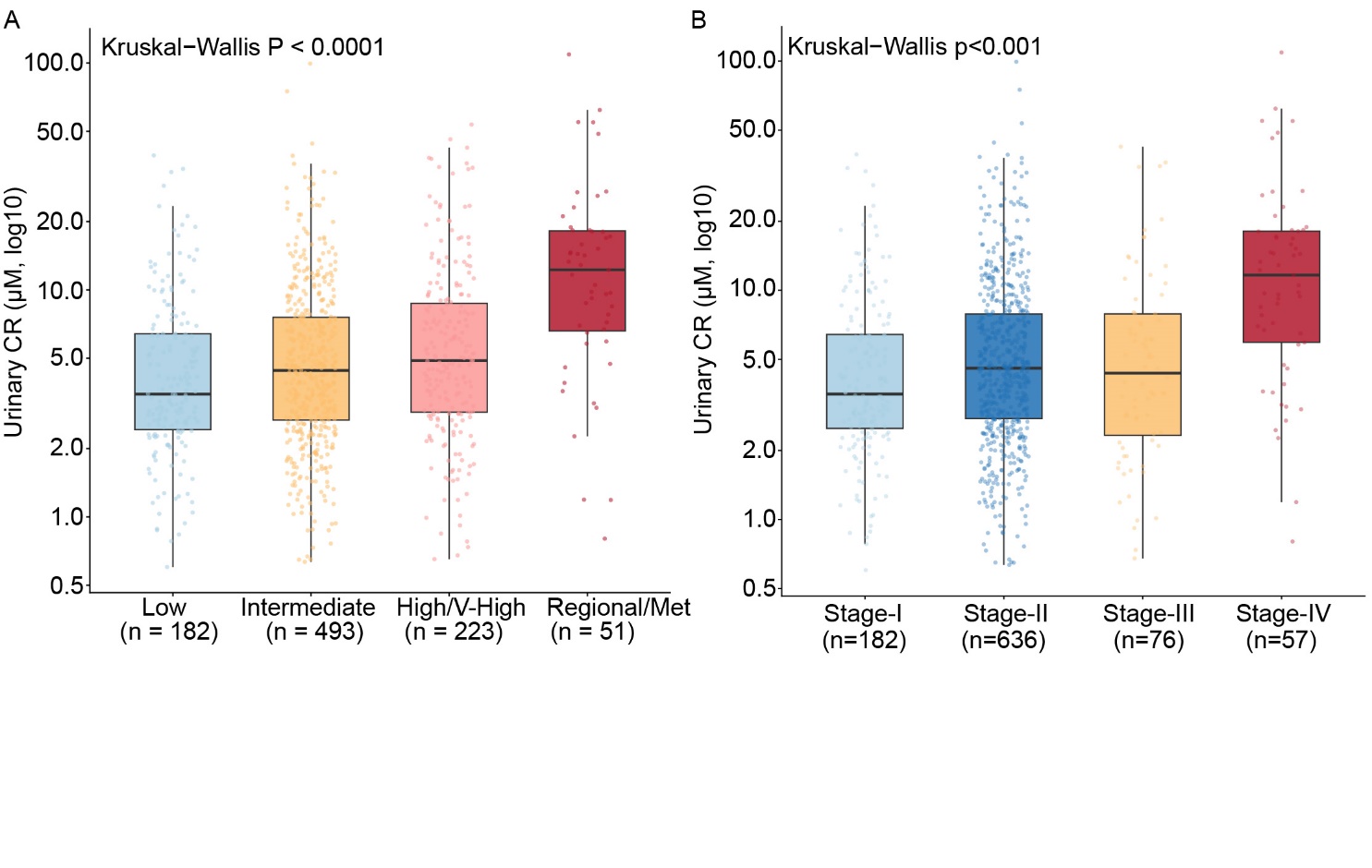

(A) Distribution of urinary CR concentrations (µM, log_10_ scale) by NCCN risk category at diagnosis (low, n=182; intermediate, n=493; high/very high, n=223; regional/metastatic, n=51), compared across groups by Kruskal–Wallis test. (B) Distribution of urinary CR concentrations (µM, log₁₀ scale) by AJCC clinical stage at diagnosis (Stage I, n=182; Stage II, n=636; Stage III, n=76; Stage IV, n=57), compared across groups by Kruskal–Wallis test. **Abbreviations:** AJCC, American Joint Committee on Cancer; CR, creatine riboside; NCCN, National Comprehensive Cancer Network.

**Supplementary Figure 4. Multinomial relative risk ratios (RRRs) for urinary CR across NCCN risk categories in the primary-model cohort and race-stratified analyses.**

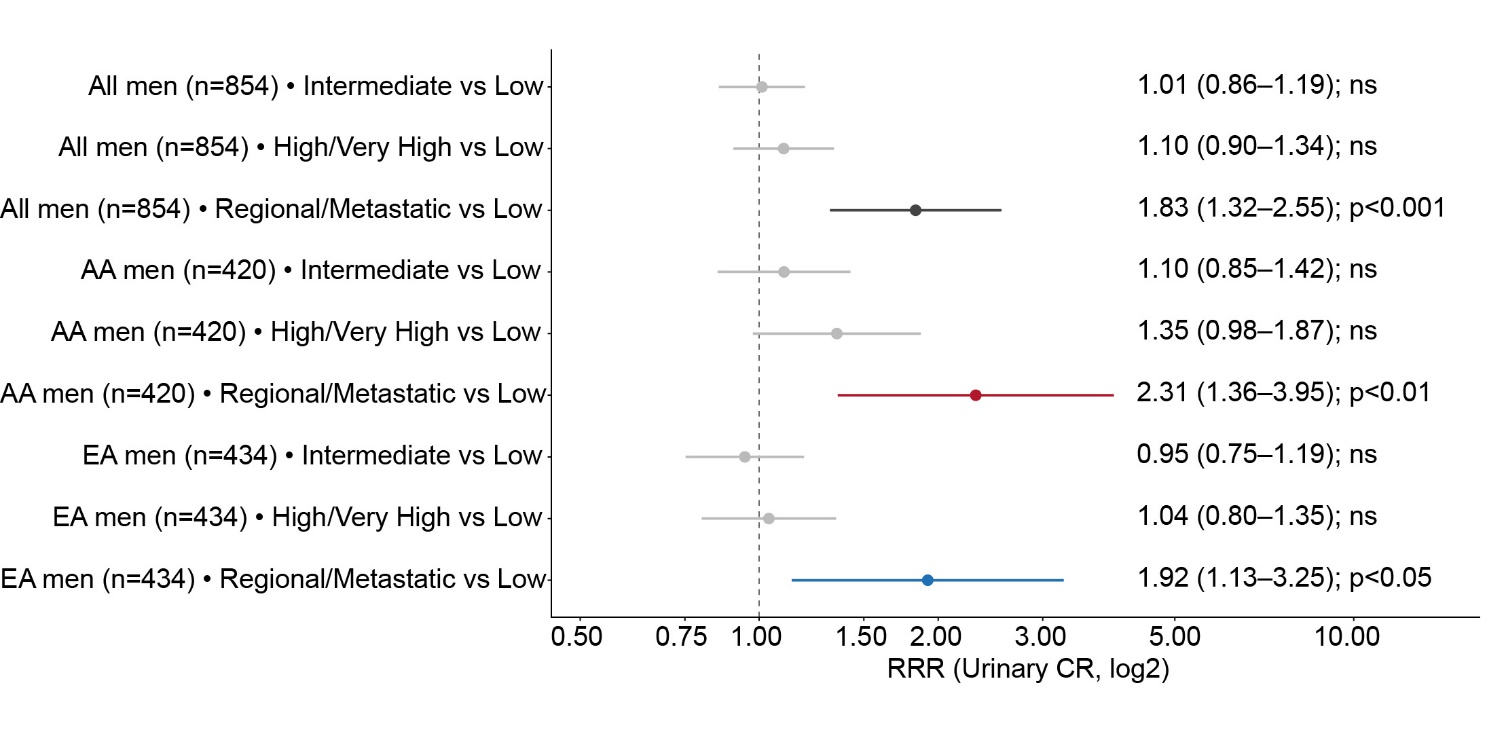

Multinomial logistic regression models showing adjusted relative risk ratios (RRR) for membership in each NCCN risk category (intermediate, high/very high, regional/metastatic) versus the reference category (low risk) per 2-fold (log₂) increase in urinary CR. Analyses are presented in the primary-model cohort (all men, n=854) and stratified by self-reported race (AA men, n=420; EA men, n=434). All models were adjusted for age, BMI, treatment modality, family history of prostate cancer, smoking status, aspirin use, diabetes, education, and individual household income. Race was not included as a covariate in race-stratified analyses. Abbreviations: AA, African American; BMI, body mass index; CI, confidence interval; CR, creatine riboside; EA, European American; ns, not significant; NCCN, National Comprehensive Cancer Network; RRR, relative risk ratio.

**Supplementary Figure 5. Stage-stratified hazard ratios for urinary CR on prostate cancer–specific mortality (n=941).**

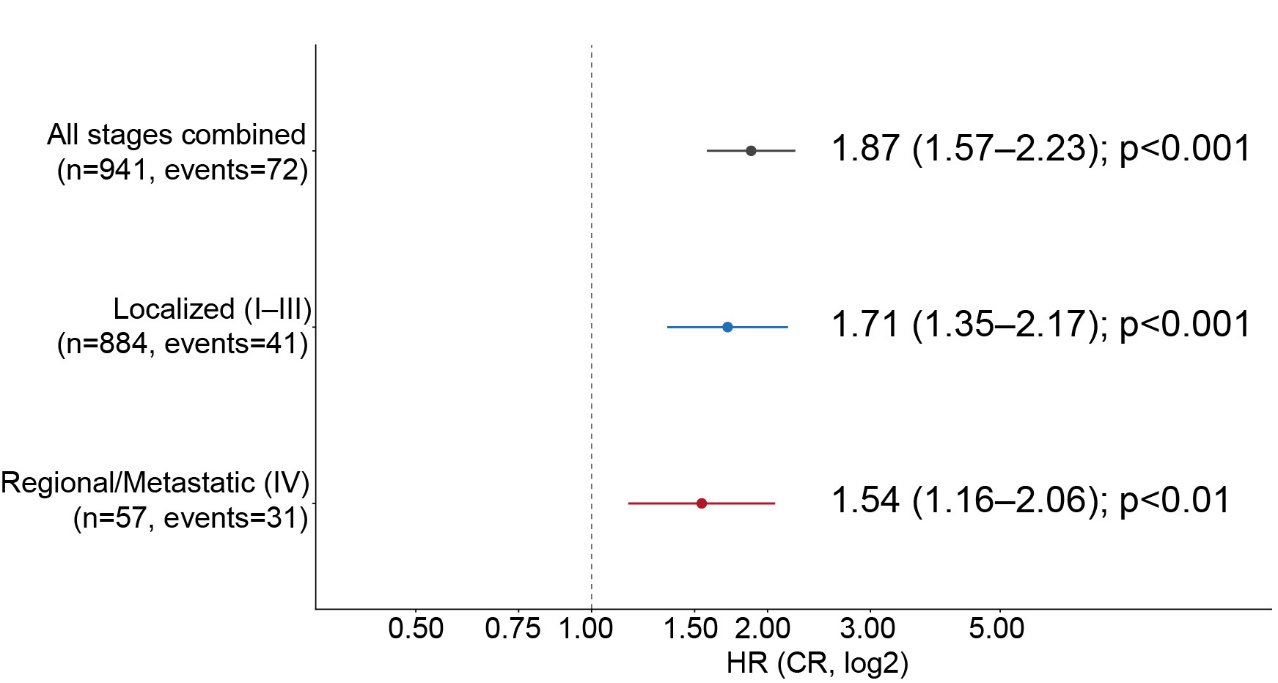

Adjusted hazard ratios for prostate cancer–specific mortality per 2-fold (log₂) increase in urinary CR, estimated using Cox proportional hazards models stratified by AJCC clinical stage at diagnosis (all stages combined, n=941 with 72 events; localized disease [Stage I–III], n=884 with 41 events; regional/metastatic disease [Stage IV], n=57 with 31 events). Parsimonious models were adjusted for age, BMI, and treatment modality. Stages IIA and IIB were combined into Stage II. Blue = localized (Stage I–III); red = regional/metastatic (Stage IV); gray = all stages combined. Abbreviations: AJCC, American Joint Committee on Cancer; BMI, body mass index; CI, confidence interval; CR, creatine riboside; HR, hazard ratio; PCa, prostate cancer.

### References

[1] Pichardo MS, Minas TZ, Pichardo CM, Bailey-Whyte M, Tang W, Dorsey TH, et al. Association of Neighborhood Deprivation With Prostate Cancer and Immune Markers in African American and European American Men. JAMA Netw Open. 2023;6:e2251745.

[2] Smith CJ, Dorsey TH, Tang W, Jordan SV, Loffredo CA, Ambs S. Aspirin Use Reduces the Risk of Aggressive Prostate Cancer and Disease Recurrence in African-American Men. Cancer Epidemiol Biomarkers Prev. 2017;26:845–53.

[3] Mohler JL, Antonarakis ES, Armstrong AJ, D'Amico AV, Davis BJ, Dorff T, et al. Prostate Cancer, Version 2.2019, NCCN Clinical Practice Guidelines in Oncology. J Natl Compr Canc Netw. 2019;17:479–505.

[4] Patel DP, Pauly GT, Tada T, Parker AL, Toulabi L, Kanke Y, et al. Improved detection and precise relative quantification of the urinary cancer metabolite biomarkers - Creatine riboside, creatinine riboside, creatine and creatinine by UPLC-ESI-MS/MS: Application to the NCI-Maryland cohort population controls and lung cancer cases. J Pharm Biomed Anal. 2020;191:113596.

[5] U.S. Food and Drug Administration. M10 Bioanalytical Method Validation and Study Sample Analysis: Guidance for Industry. Silver Spring, MD: Food and Drug Administration; 2022.
